# Can diffuse reflectance spectroscopy identify shuntodynia in pediatric hydrocephalus patients?

**DOI:** 10.1101/2023.10.17.23297150

**Authors:** Olivia Kline, Karthik Vishwanath, Boyd Colbrunn, Andrew Peachman, Jing Zhang, Sudhakar Vadivelu

## Abstract

**Significance:** Shuntodynia is patient reported pain at the site of the implanted ventriculo-peritoneal (VP) shunt. Pediatric hydrocephalus requiring shunt placement is a chronic and prevalent standard of care treatment and requiring lifetime management. Shuntodynia is a subjective measure of shunt dysfunction. Quantitative, white-light tissue spectroscopy could be used to objectively identify this condition in the clinic.

**Aim:** Pediatric subjects were recruited for optical sensing during routine clinical follow-up visits, post VP shunt implantations. Acquired optical signals were translated into skin-hemodynamic signatures and were compared between subjects that reported shuntodynia vs. those that did not.

**Approach:** Diffuse reflectance spectroscopy (DRS) measurements were collected between 450-700 nm using a single-channel fiber-optical probe from (N=35) patients. Multiple reflectance spectra were obtained by the attending physician from regions both proximal and distal to the VP shunt sites, and from a matched contralateral site for each subject. Acquired reflectance spectra were processed quantitatively into functional tissue optical endpoints. A two-way, repeated measures analysis of variance (ANOVA) was used to assess whether and which of the optical variables were statistically separable, across subjects with shuntodynia vs. those without.

**Results:** Results showed that vascular oxygen saturation was significantly lower in subjects reporting shuntodynia, when measured proximal to shunt sites. Subjects with shuntodynia also had lower total hemoglobin at the shunt site relative to distal sites. Both patient groups showed higher tissue scattering at the shunt sites in comparison to the contralateral sites.

**Conclusions:** Optically derived hemodynamic variables were statistically significantly different in subjects presenting with shuntodynia relative to those without. DRS could provide a viable mode in routine bedside monitoring of subjects with VP shunts for clinical management and risk assessment of shuntodynia.

## 1 Introduction

Hydrocephalus is characterized by an increased intracranial cerebrospinal fluid (CSF) volume that is independent of hydrostatic or barometric pressure ^1^. This condition is prevalent and costly with some sources estimating that hydrocephalus costs the United States healthcare system nearly two billion dollars annually ^2, 3^. Hydrocephalus can be a result of several ailments including tumors, degenerative diseases, or trauma and may occur at any age and is also known to arise congenitally in pediatric patients (with one case of hydrocephalus for every 1000 births) ^1, 4^. With a diverse range of associated symptoms, hydrocephalus reduces the quality of life for child-patients during diagnostic evaluation and possibly even after treatment .

Ventriculoperitoneal (VP) shunts are the gold standard in treating pediatric hydrocephalus ^5^. VP shunts however elicit challenges for patients including infection, mechanical and functional issues ^6^. Additionally, nearly half of pediatric hydrocephalus patients reported headaches and generalized pain after VP shunt placements ^7^. Here, we use the term shuntodynia to indicate patient-reported pain that is reported clinically in patients with an implanted VP shunt. Specifically, this pain can occur at the shunt valve site or more distally along the shunt distal tubing. Interestingly, shuntodynia has been reported with and without shunt malfunctions ^8, 9^. Though clinically recognized, the condition has not received significant attention since it is usually seldom life-threatening. The presence of shuntodynia is indicative of patient discomfort, and there is significant physical and psychosocial burden associated with implanted shunt devices ^10^.

Optical sensing techniques such as near infrared spectroscopy (NIRS) are well suited to explore questions pertaining to pain since they are noninvasive, painless and can be acquired during noxious stimuli ^11^. The use of such methods for objective assessment of pain has widely been explored through measurement of hemodynamic responses in various brain regions via functional NIRS ^12-15^. Optical spectroscopic methods have also been widely used to monitor and assess pain in exercise science and sports-medicine ^16-19^. Parallelly, a related diffuse optical technique called diffusion correlation spectroscopy (DCS) has been used in pediatric subjects with hydrocephalus for rapid and non-invasive assessment of intracranial pressure (ICP)^20-24^. However, to the best of our knowledge optical sensing has not been applied or explored in the realm of shuntodynia this far. Additionally, most previous studies that have used optical sensing for pain studies have mostly focused on using these methods for detecting functional changes (such as blood oxygenation, volume or tissue perfusion) from deep tissue layers. Again, to the best of our knowledge no previous diffuse optical studies have examined optically derived functional changes in skin tissues. We have previously employed fiber-based diffuse reflectance spectroscopy (DRS) systems operating at short source-detector geometries to quantify functional properties in skin tissues *in vivo* ^25-27^. We have also previously reported preliminary findings from our use of DRS in pediatric patients implanted with VP shunts for hydrocephalus with or without shuntodynia ^28^. Here, we extend our analysis to be statistically robust and examine if patient reported shuntodynia was detectable using optically derived, quantitative hemodynamic signatures obtained in skin-tissues adjacent to VP implants in subjects, during a single clinical visit.

## 2 Methods

### 2.1 Study protocol and clinical data collection

All procedures for data collection were conducted with approval from the Institutional Review Board (IRB) at Cincinnati Children’s Hospital Medical Center (CCHMC). Informed consent was obtained from the patient’s parent or legal guardian during routine clinic appointments. Invitation to participate in the study was offered to pediatric hydrocephalus patients that had a VP shunt, at the discretion of the attending neurosurgeon. Patients with an inability to stay still for duration of DRS scans (which ranged to between 5-15 minutes across subjects) or those that presented hypersensitivity to placement of optical probe were excluded. Of the 35 patients (N=9 with shuntodynia) that were recruited in the study, data from 4 subjects was missing (due to computer glitches), while one subject had VP shunts on both sides and was excluded. 30 subjects (N=8 with shuntodynia) had complete datasets and were used for the analysis reported here.

Optical data collection was performed by having the patients in the seated position. DRS scans were collected at 7 different spatial sites for N=25 subjects (4 sites circling the shunt labeled by o’clock positions, 2 at the scar sites proximal and distal to shunt and 1 from a matched contralateral site on the opposite side to where the shunt was implanted) as a within-subjects control. For 5 subjects, data from the contralateral sites were not collected.

### 2.2 Instrumentation

DRS measurements were obtained using a fiber spectroscopy system described previously ^25, 28^. The system was constructed by coupling a broadband tungsten-halogen light source (HL2000, Ocean Optics, FL) to the tissue using a bifurcated optical fiber with core size 600 μm and collecting the diffuse reflectance at a distance of approximately 600 μm from the source, with an identical fiber (BIF600-UV-VIS, Ocean Optics, FL). The average depth sensed under the probe was not expected to exceed 300-500 μm with these probe dimensions and optical spectrum. The detected reflectance was directed to the entrance slit of a spectrometer (USB4000, Ocean Optics, FL) for collection. On each day, prior to data collection from the patient, baseline reference spectra were collected from a white spectralon reflectance standard (Labsphere, North Sutton, NH, USA) and from a homogeneous solid reference phantom with known optical properties (INO, Hamilton, Ontario, Canada) for calibration as described before ^26, 29^. Integration times on the spectrometer for collection of a single DRS scan collection ranged between 10-35 ms. For each subject, dark scans were also obtained with the probe placed on subjects but with lamp shuttered. For each tissue site, five consecutive scans were collected and stored.

### 2.3 Data fitting and filtering

Optical measurements acquired from subjects were analyzed using MATLAB (Mathworks, Natick, MA). Each spectrum was corrected for spectrometer integration time, dark-noise subtracted and then normalized by the spectrum of the white reflectance standard, giving a DRS spectrum that could be inverse-fitted to extract wavelength absorption and reduced scattering coefficients using a previously reported Monte Carlo (MC) based photon transport model ^27, 29^. The absorption coefficient was translated into oxygenated and deoxygenated hemoglobin molar concentrations ([HbO2] and [dHb]) from published molar extinction coefficients of these chromophores and added to yield the total hemoglobin concentration ([THb].) Vascular oxygen saturation was calculated as a ratio of oxygenated hemoglobin to total hemoglobin and reported as a percent value where SO_2_=100 × [HbO_2_]/[THb]. Finally, a wavelength–averaged scattering coefficient was calculated using the wavelength-dependent scattering from the inverse MC model which assumed medium anisotropy of 0.9 with the Henyey-Greenstein phase function ^29^.

For each DRS that was fitted by the inverse model, a goodness of (χ^2^) was calculated as

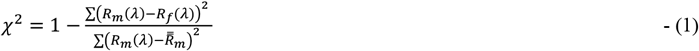

In equation (1), *R*_*m*_(λ) and *R*_*f*_(λ) are measured and fitted DRS, respectively with the sum computed across all wavelengths and 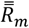 was the wavelength-averaged measured reflectance. We excluded data with χ^2^ < 0.9 from statistical analysis. This resulted in a loss of 28 DRS scans (10.4%) from the pain group and 69 scans (8.9%) from the no-pain group. After filtering the data based on this condition, multiple scans for shunt and scar sites for each patient remained. However, this process resulted in a reduction of contralateral scans available for statistical analysis (with 3 subjects from the pain group and 2 from no-pain group not having any contralateral site information).

### 2.4 Statistical modeling

Optically derived values of vascular oxygen saturation (SO_2_), total hemoglobin ([THb]), and wavelength-averaged reduced scattering (< µ_*s*_ >) from each DRS scan (across all subjects and sites) were analyzed using repeated measures analysis of variance (ANOVA). These models were performed using “lme4” package in R ^30^. To adjust the p-value of simultaneous inference, we conducted Tukey p-value adjustment in the post-hoc analysis via “emmeans” package in R ^31^. These statistical techniques were selected to evaluate whether optical data were influenced by the presence (lr lack) of shuntodynia at different sites. Furthermore, it also incorporated correlations present in repeated measurements obtained from the same subject.

Statistical comparisons were first made to identify if derived coefficients at different tissue sites within the same patient subset were separable (e.g., shunt to contralateral comparisons for all patients with shuntodynia). Comparisons were then also made across patient groups to compare site-level responses measured (e.g., pain vs, no pain at the shunt site). Results are presented from the two-way repeated measures ANOVA.

## 3 Results

### 3.1 Optical Data

Fig. 1 shows the averaged DRS data collected from all subjects enrolled in the study based on the reported pain status and tissue sites (Fig. 1a-1c for sites around the shunt, the scar tissue and a matched contralateral region, respectively). The mean diffuse reflectance observed at both the shunt and scar sites, the shuntodynia group to have higher reflectance (Fig. 1a and 1b) which was not seen at the contralateral sites. Additionally, the variance across subjects was higher in the shuntodynia group in comparison to the no-pain group.

**Fig. 1.**
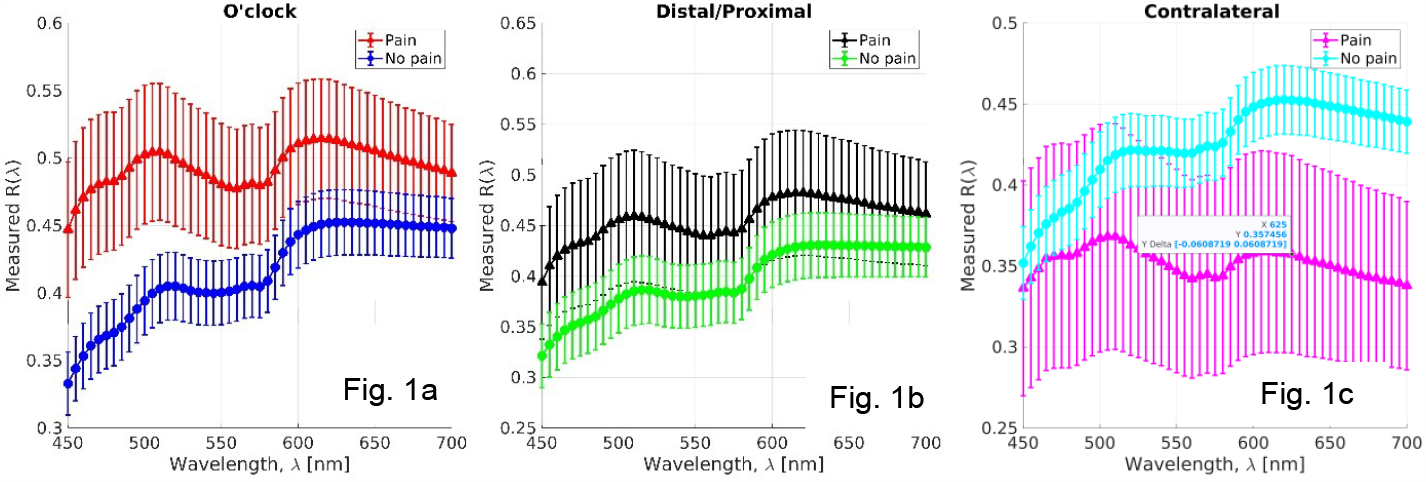
Averaged reflectance data across all patients in the pain and no-pain groups obtained at the three different sites probed (a) DRS data for all shunt sites, (b) DRS data for all scar sites, and (c) DRS data on a contralateral side matched to the shunt site. Each figure shows average DRS for each patient group (symbols) while the error bars show standard deviations.

Each measured DRS spectrum was fitted and filtered for goodness of fits as in equation (1). Figure 2 shows the calibrated DRS measurements from two representative subjects (Fig. 2a, 2c and 2e for a subject with shuntodynia and Fig. 2b, 2d and 2f for a subject without), for each site when fitted by the inverse MC model. Symbols indicate measured values while lines indicate MC fits and all data shown here had χ^2^ values between 0.9 and 0.92. Fig. 2c and 2d show the extracted absorption coefficients for the subject in pain and no-pain group, respectively (Fig. 2e and 2f show the derived scattering coefficients). These optical coefficients were obtained from the inverse MC model as a result of fitting the data shown in Fig. 2a and 2b, respectively^26, 29^. As seen visually, the measured data were well fit by the MC model and the extracted absorption spectrum showed characteristic features between 550-600 nm for oxygenated hemoglobin. The scattering coefficients were derived in the MC model by assuming that the anisotropy was fixed at 0.9 for the medium and by modeling the scattering as a power law, µ_*s*_ = *A*λ^−*b*^ and estimating *A, b* in the optimization process.

**Fig. 2.**
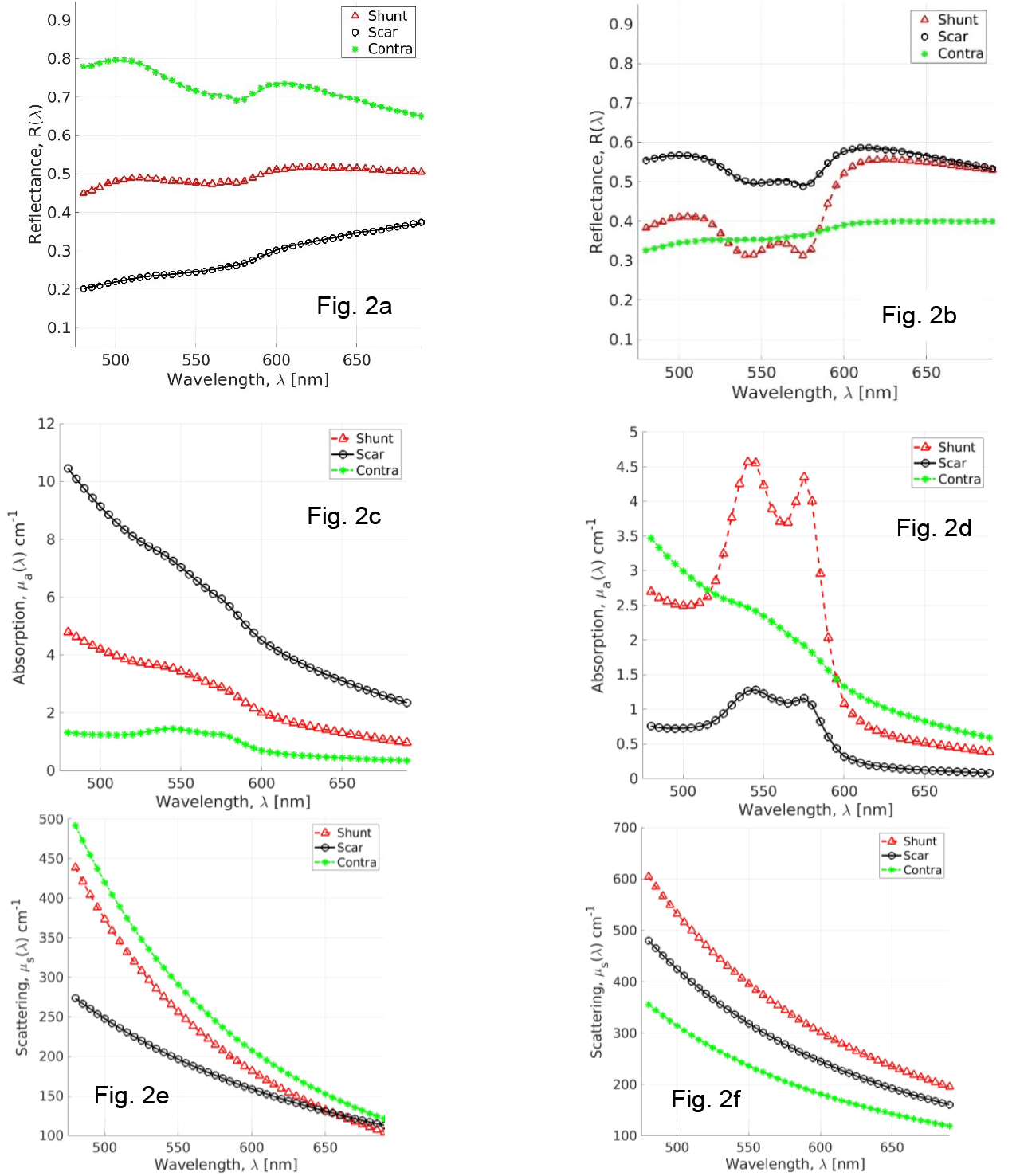
Measured (symbols) and fitted (lines) DRS data from two representative subjects (Fig. 2a: with shuntodynia; Fig. 2b: without shuntodynia). Fig. 2c and 2e show derived absorption and scattering coefficients for each DRS data fitted in Fig 2a while Fig. 2d and 2e show these data for data shown in Fig. 2b

### 3.2 Hemodynamic Data

The absorption coefficients from inversely fitted DRS spectra were translated into the traditionally used optically derived functional endpoints of total hemoglobin ([THb] = [HbO_2_]+[dHb]) and vascular saturation percent (SO_2_ = 100×[HbO_2_]/THb]), while the scattering coefficient was averaged across the spectrum and multiplied by (1-*g*) to get the average reduced scattering (< µ_*s*_′ >). Thus, each DRS spectrum was reduced into three quantitative optical coefficients which were analyzed statistically. Fig. 3 shows a comparison of these three optically derived funcrional variables, at each tissue site, for both patient groups. The mean oxygen saturation and total hemoglobin values appeared to be similar between the shunt-scar sites for the patients that did not present with shuntodynia but those with shuntodynia had elevated total hemoglobin and oxygen saturation in the scar sites relative to the shunt sites.

**Fig. 3.**
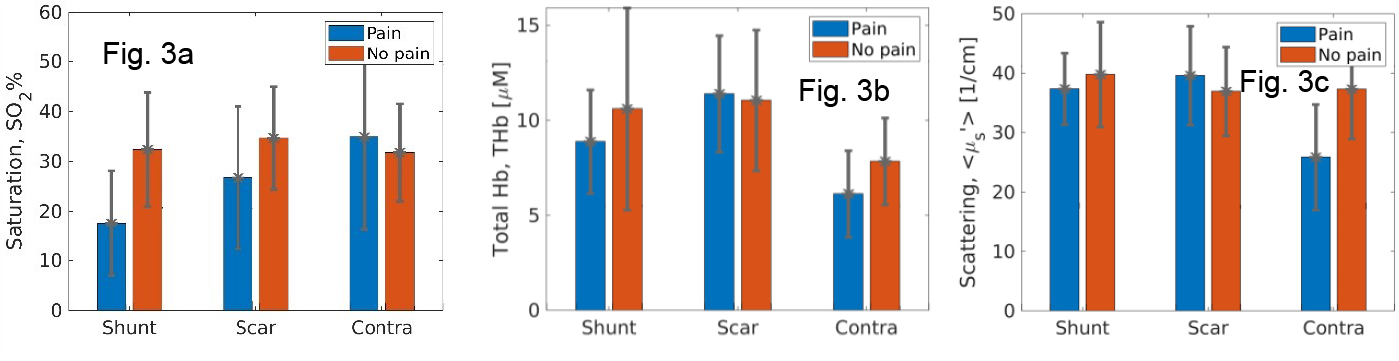
Comparison of mean optically derived hemodynamic and scattering coefficients across the three tissue sites for each patient group (blue bars: with shuntodynia; red bars: without shuntodynia). Bars represent mean value while error bars depict standard deviations. Data in Fig. 3a show the vascular oxygen saturation, in 3b are for total hemoglobin concentration and Fig. 3c are for the mean wavelength averaged reduced scattering.

Table 1 presents a summary of these results along with statistical analysis obtained for site- wise comparisons in each patient group. Table 1 lists means and standard errors for optical variables at each site, for both patient groups. Tukey-adjusted p-values are shown also shown for comparing each corresponding optical variable measured at two distinct tissue sites. Comparisons were made within the pain and no pain groups with the intention of establishing optically observed differences at different sites, within individual patients. Statistical analysis was not performed to establish a difference between the pain and no pain groups at the same site because the goal of this preliminary study was to try and seek optically derived metrics that could help with identifying the presence of shuntodynia within any individual patient.

**Table 1.** Tukey-adjusted, pairwise comparisons of optical variables within patient groups with and without shuntodynia. Means and standard errors for the groups are shown as organized by site and optical variable. P-values are obtained from adjusted Tukey comparisons to estimate if mean differences in relevant variable-pairs across different sites was zero or not. NS indicates p-values above 0.05.

| Patient Group | Optical variable | Mean ± Std values at | | | p-values for site-site differences | | |
| --- | --- | --- | --- | --- | --- | --- | --- |
|  |  | Shunt | Scar | Contra-lateral | Shunt vs. scar | Shunt vs. Contra | Scar vs. Contra |
| Shuntodynia | SO<sub>2</sub> (%) | 17.9±4.0 | 24.9±4.3 | 35.4±5.6 | <0.05 | <0.01 | NS |
| | [THb] μM | 8.7±1.6 | 11.2±1.7 | 7.0±2.1 | 0.05 | NS | <0.05 |
| | < $\mu_s$ > cm<sup>-1</sup> | 213±16 | 220±18 | 143±23 | NS | <0.01 | <0.01 |
| No shuntodynia | SO<sub>2</sub> (%) | 33.0±2.4 | 35.4±2.7 | 32.3±3.0 | NS | NS | NS |
| | [THb] μM | 10.5±1.0 | 11.0±1.0 | 7.5±1.2 | NS | <0.01 | <0.01 |
| | < $\mu_s$ > cm<sup>-1</sup> | 222±10 | 208±11 | 205±12 | NS | NS | NS |

Oxygen saturation proved to be the most useful metric in identifying pain within the group presenting with shuntodynia. For the pain group, oxygen saturation was significantly lower than both the scar and control sites. The extracted total hemoglobin coefficient also exhibited statistical significance within the shuntodynia group where this variable had elevated values at the scar site relative to the shunts sites. Finally, the control contralateral site showed significantly lowered scattering coefficient than all other sites for the patients experiencing pain and may be a statistical anomaly given the low number of contralateral scans available for this patient group.

Interestingly, in patient group without shuntodynia, oxygen saturation was not significantly different across any of the measured tissue sites. Likewise, the scattering coefficient also did not indicate any statistically significant differences between sites. However, total hemoglobin was elevated at both scar and shunt sites which were located ipsilaterally on the patient’s heads as the implanted shunts. Though statistical comparisons were not performed between patient groups at the same site, Figure 3 appears to indicate that the two patient groups could show differences at certain sites for some of these optical variables.

## 4. Discussion

Fiber based DRS was used in a preliminary clinical study to objectively measure the hemodynamic status of skin tissue in pediatric patients that had implanted VP shunts as a part of their treatments for hydrocephalus. The population studied here either presented with idiopathic clinical pain around the VP shunt and were placed in the shuntodynia group while the others were placed into the no-shuntodynia group. The optically acquired measurements were translated into three functional endpoints of vascular oxygen saturation, the total hemoglobin concentration, and the wavelength averaged tissue-scattering. These optical endpoints were then analyzed using a two-way ANOVA to statistically model the presence of shuntodynia and address the question of whether shuntodynia could be detected in individual subjects by looking across different tissue sites. For each subject, optical measurements were acquired from three different tissue sites as described above. To the best of our knowledge, this work marks a first attempt to use quantitative DRS to extract optically derived functional metrics from skin tissue surrounding implanted VP shunts for the objective detection of shuntodynia.

Statistical analysis showed that patients with the condition had lower oxygen saturations than patients without pain at the shunt sites. Additionally, patients with pain exhibited lower oxygen saturations at the shunt and scar sites than was measured from a matched contralateral side. In comparing the shunt sites for both groups, we observed that the average oxygen saturation for subjects with pain was lower. Similarly, in comparing sites for pain patients, we saw that shunt SO_2_ was lower than measurements from scar or the contralateral sites. However, since our sample size in the pain group was small, the variations in measurements were larger. The most significant finding was that a decreased oxygen saturation was associated at the shunt site, in subjects that complained of shuntodynia.

With a confidence interval of 95%, it was not possible to identify a difference in total hemoglobin for patients who had pain and those who did not. However, in patients without pain, our results indicated that total hemoglobin was higher at the shunt and the scar sites than it was in the contralateral side. In patients presenting with shuntodynia, total hemoglobin was significantly higher at the scar sites than it was at the shunt or contralateral sites. For the no-shuntodynia group, we observed greater variability in acquired measurements at the shunt and scar sites. Overall, there was no discernable difference in the mean scattering coefficients for patients with and without shuntodynia. Although, we did observe a statistically significant difference for scattering measurements in the pain group, where the contralateral side exhibited lower scattering coefficient relative to both scar and shunt sites, given the low number of scans available for contralateral measurements this conclusion remains to be validated in future studies.

Of the three optical variables derived from experimental measurements of DRS, oxygen saturation presented as the most promising metric for identifying shuntodynia. Not only was there a distinction between the pain and no pain groups for oxygen saturation, but pain patients exhibited a lower oxygen saturation around their shunt sites. This finding could also be supported by reported assertions of oxygen saturation being a potential biomarker of pain, whether directly related to increased sympathetic activity versus hypovolemia ^32^.

## 5 Conclusion

Here, we present preliminary findings that suggest there were differences in optically estimated hemodynamics within skin tissues of pediatric hydrocephalus patients who experienced shuntodynia post VP shunt implantations. Our results showed that oxygen saturation presented the greatest statistical value for identifying the presence of pain called shuntodynia. Though the use of DRS was direct and non-invasive, the fiber-based system required contact with skin tissue. As a practical issue in this study, the clinical condition we studied naturally presented impediments to establish good contact between the probe and skin tissues in subjects. These took significant effort and costs in terms of clinical time and patient management for appropriate optical data collection. Thus, having fully non-contact optical methods for sensing tissue hemodynamics would vastly improve data collection ability, especially when working with patients experiencing pain. Additionally, it will also be valuable to integrate optical tissue perfusion measures such as diffusion correlation spectroscopy to build a more complete picture of subcutaneous hemodynamics in shuntodynia.

## Disclosures

The authors declare that there are no conflicts of interest related to this article.

## Data Availability

All data produced in the present study are available upon reasonable request to the authors

## Acknowledgments

We acknowledge the assistance of clinical faculty and staff at Cincinnati Children’s Hospital Medical Center and thank them for helping make their work possible. We also acknowledge support from intramural funds at Miami University (Undergraduate Summer Scholar to BC and OK, and the Undergraduate Research Award to OK).

## Code, Data, and Materials Availability

Code used to collect and analyze data was developed in-house. Programs were written to facilitate the process of obtaining scans and also inverting them via the inverse Monte-Carlo model.

## Notes

### Competing Interest Statement

The authors have declared no competing interest.

### Funding Statement

This study did not receive any extramural funding

### Author Declarations

The IRB of Cincinnati Childrens Hospital Medical Center gave ethical approval for this work

